# Genetic basis of dynamic brain states reveals cellular and disease associations

**DOI:** 10.64898/2026.06.10.26355409

**Authors:** Amir Ebneabbasi, David J Whiteside, Yuanjun Gu, Richard A.I. Bethlehem, Varun Warrier, Timothy Rittman

## Abstract

Dynamic resting-state fMRI captures the time-varying patterns of brain activity that are obscured by static approaches. Hidden Markov Models (HMMs) characterise these dynamics as recurring whole-brain states and quantify their fractional occupancy (FO), the proportion of time spent in each state, yet the biological basis of inter-individual variation in FO remains unclear. Using data from 52,335 White UK Biobank participants, with replication in East and South Asian subsamples, this study examined the heritability, cellular and neurotransmitter basis of brain states, and their links with complex phenotypes. FO was significantly heritable and enriched for neuronal populations, particularly glutamatergic and GABAergic signalling. Analyses identified shared and state-specific loci and revealed genetic correlations, colocalisation, and potential causal relationships between FO and several phenotypes, including educational attainment, sleep duration, and disease risk. These findings establish dynamic brain states as biologically grounded intermediate phenotypes, linking genetic variation to neural dynamics, diseases and traits.

## Introduction

The human brain is characterised by continuous and spontaneous activity, which emerges independently of deliberate task performance or external stimulation. Resting-state functional magnetic resonance imaging (fMRI) has demonstrated that brain regions do not operate independently; instead, they form functionally connected networks whose activity is temporally correlated [1, 2]. While helpful in mapping networks, static measures cannot capture the time-varying nature of brain function. Instead, there is a growing understanding that brain activity is inherently dynamic, transitioning between recurring whole-brain configurations over time [3, 4].

Hidden Markov Models (HMMs) have emerged as a robust framework for identifying discrete *brain states* from fMRI data. By modelling each state as a multivariate Gaussian distribution, the HMM approach captures the state-specific temporal evolution of both activation and connectivity [5, 6]. This avoids many of the arbitrary parameter choices and statistical limitations associated with sliding-window methods. Notably, HMMs offer interpretable temporal properties, including state-specific fractional occupancy (FO), which quantifies the proportion of time the brain spends in each state and therefore provides a concise measure of inter-individual variability. Interestingly, FO is widely linked to cognitive function [7, 8], ageing trajectories [9], and neuropsychiatric conditions [10, 11]. Despite these phenotypic associations, the extent to which genetic factors shape the FO of brain states—and their potential relevance to disease—remains unclear.

The present study focuses on three main goals. First, we conduct a genome-wide association study (GWAS) of state-specific FO in a cohort of European participants (n=52,335), followed by replication in two subsamples of East and South Asians. This complements prior GWASs on static fMRI features [12–19] and leverages the largest sample size to date for studying dynamic connectivity, enabling a robust interrogation of the heritability of brain states. We employ multiple gene-mapping approaches and evaluate enrichment across specific cell types and neurotransmitter systems, thereby providing translational insight into the biological substrates that shape brain dynamics.

Second, although extensive pleiotropy—where a single genetic variant influences multiple traits—has been observed both within brain regions for individual imaging-derived phenotypes (IDPs) and across different IDPs [12–20], it remains unclear whether the distinct activation and connectivity patterns captured by HMM-derived brain states reflect unique genetic influences or a shared set of genetic factors acting differentially across states. Clarifying this distinction is essential for interpreting the biological validity of brain states. We address this by interrogating the genetic architecture of brain states for pleiotropy.

Finally, we aim to examine whether the genetic architecture of brain states overlaps with traits and diseases. We assess genome-wide genetic correlations, which quantify the shared polygenic contribution between phenotypes, and perform colocalisation analyses to determine whether overlapping association signals at specific loci reflect the same underlying causal variant. In addition, bidirectional causal relationships are explored using Mendelian randomisation, which leverages genetic variants as instrumental variables to infer directionality. Together, these complementary approaches clarify how polygenic risk for health and disease may act through dynamic neural processes, positioning brain states as potential intermediates between genes and clinical outcomes.

## Results

Denoised resting-state fMRI data from 52,335 white British participants were used in our study [21, 22]. We leveraged the UK Biobank group ICA results, selected 55 non-artefactual components from the original 100, and used the associated subject-specific time series for subsequent analyses. We then applied HMM to segment each participant’s data into 12 recurrent states. Each state is characterised by a mean activation vector and a connectivity covariance matrix [5, 6]. Model estimation was performed using variational Bayes, optimised for big data with a stochastic algorithm (see Methods).

The activation patterns of the 12 brain states (Fig. 1) and their statistical distributions are shown (Extended Data Fig. 1). To clarify the spatial configuration of the brain states, we projected activation maps onto seven well-recognised functional connectivity networks [23]. For this purpose, group-level activation maps were first parcellated into 180 bilaterally averaged regions [24], after which correlations with network categories were computed while controlling for spatial autocorrelation [25] (Extended Data Fig. 2). Overall, states 1, 8, and 11 represented the default mode network, and states 3, 6, 7, 9, and 10 were associated with the visual and sensorimotor networks. States 2, 4, and 5 were linked to the dorsal and ventral attention networks, and state 12 to the frontoparietal network. These patterns suggest partially overlapping functional roles across brain states. Next, to quantify the temporal dynamics of these states, we computed fractional occupancy (FO) from the posterior state probabilities at each time point, defined as the proportion of time each participant spent in a given state. FO measures were used as quantitative phenotypes for genome-wide association analyses.

**Fig. 1.**
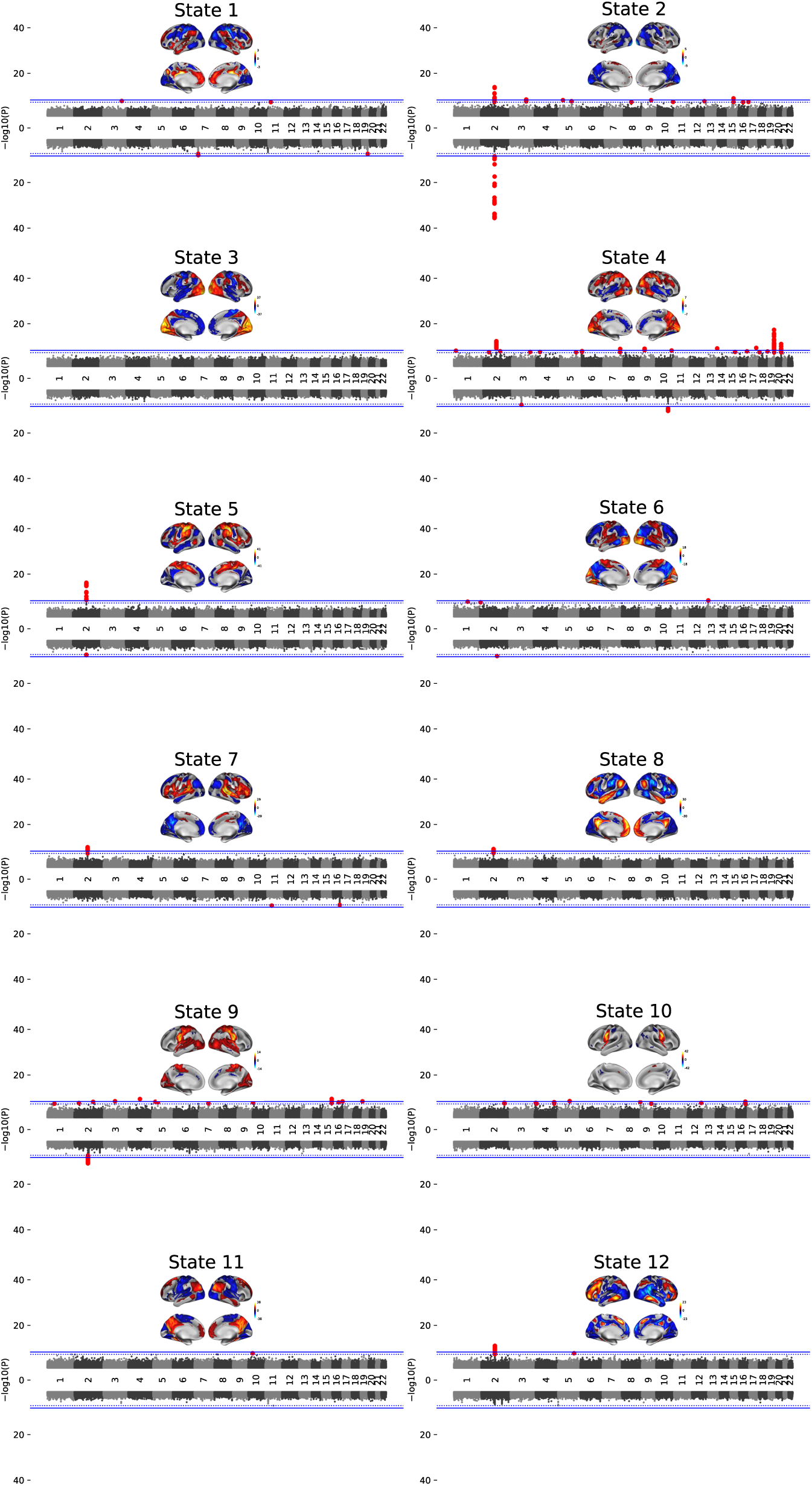
Activation patterns of 12 dynamic brain states estimated using a Hidden Markov model in the UK Biobank, alongside genome-wide association analyses of their fractional occupancy (FO) using a mixed linear model. Genetic variants exceeding the genome-wide significance threshold (p < 5 × 10⁻⁸) are highlighted in red, with the corresponding threshold indicated by a dashed line. A more stringent Bonferroni-corrected threshold (p < 5 × 10⁻⁹), accounting for the 12 states, is also shown.

SNP array data were obtained from the UK Biobank [26] and underwent additional in-house quality-control procedures (see Methods). After standard filtering for MAF > 0.0001, 19,914,312 SNPs were retained for subsequent analyses. To account for cryptic relatedness among samples, a genetic relatedness matrix (GRM) was constructed from LD–pruned SNPs (n = 565,631) [27]. Ancestral diversity was controlled for using genetic principal components (PCs) obtained via fastPCA [28]. A mixed linear model (MLM) genome-wide association study (GWAS) was then performed separately for each FO phenotype, with adjustment for covariates of no interest (Methods). Miami plots are presented (Fig. 1).

### Brain States Are Heritable, Linked to Meaningful Genes, Cell Types and Neurotransmitters

Clumping with PLINK [29] using *p* < 5 × 10⁻⁸, a 1000 kb window, and *r*² = 0.1 identified 44 independent loci, with the number of loci per state ranging from zero (State 3) to 12 (State 4) (Supplementary Table 1). The total number of genome-wide significant SNPs per state varied substantially, from seven in State 10 to 276 in State 7. To estimate SNP-based heritability (h2), we primarily used Genome Restricted Maximum Likelihood (GREML) [30] (Fig. 2A and Supplementary Table 2). GCTA-GREML estimates ranged from 0.0355 (SE = 0.0182) to 0.1332 (SE = 0.0195), with all heritability estimates being statistically significant. GCTA-GREML showed that state 9 had the highest heritability estimate (0.1332, SE = 0.0196, *p* = 9.25 × 10^-13^), followed by states 2 (0.1209, SE = 0.0194, *p* = 8.64 × 10^-11^) and 1 (0.1162, SE = 0.0192, *p* = 1.46 × 10^-10^). LDSC-based heritability [31] estimates were similar in ranking, although smaller, with state 9 again showing the highest heritability (0.0734, SE = 0.0097) (Supplementary Table 3). We examined the attenuation ratio (calculated as LDSC intercept − 1 / mean χ² − 1), which quantifies the proportion of inflation attributable to factors other than polygenic heritability. Q–Q plots are also shown to assess the calibration of GWAS association statistics (Extended Data Fig. 3). Across the 12 brain states, ratios ranged from 0.072 (SE = 0.133) to 0.317 (SE = 0.103), with the highest value in State 3, which showed no genome-wide significant loci, and the lowest in State 10. Overall, low ratios indicate that inflation is primarily due to polygenicity rather than confounding (Supplementary Table 4).

**Fig. 2.**
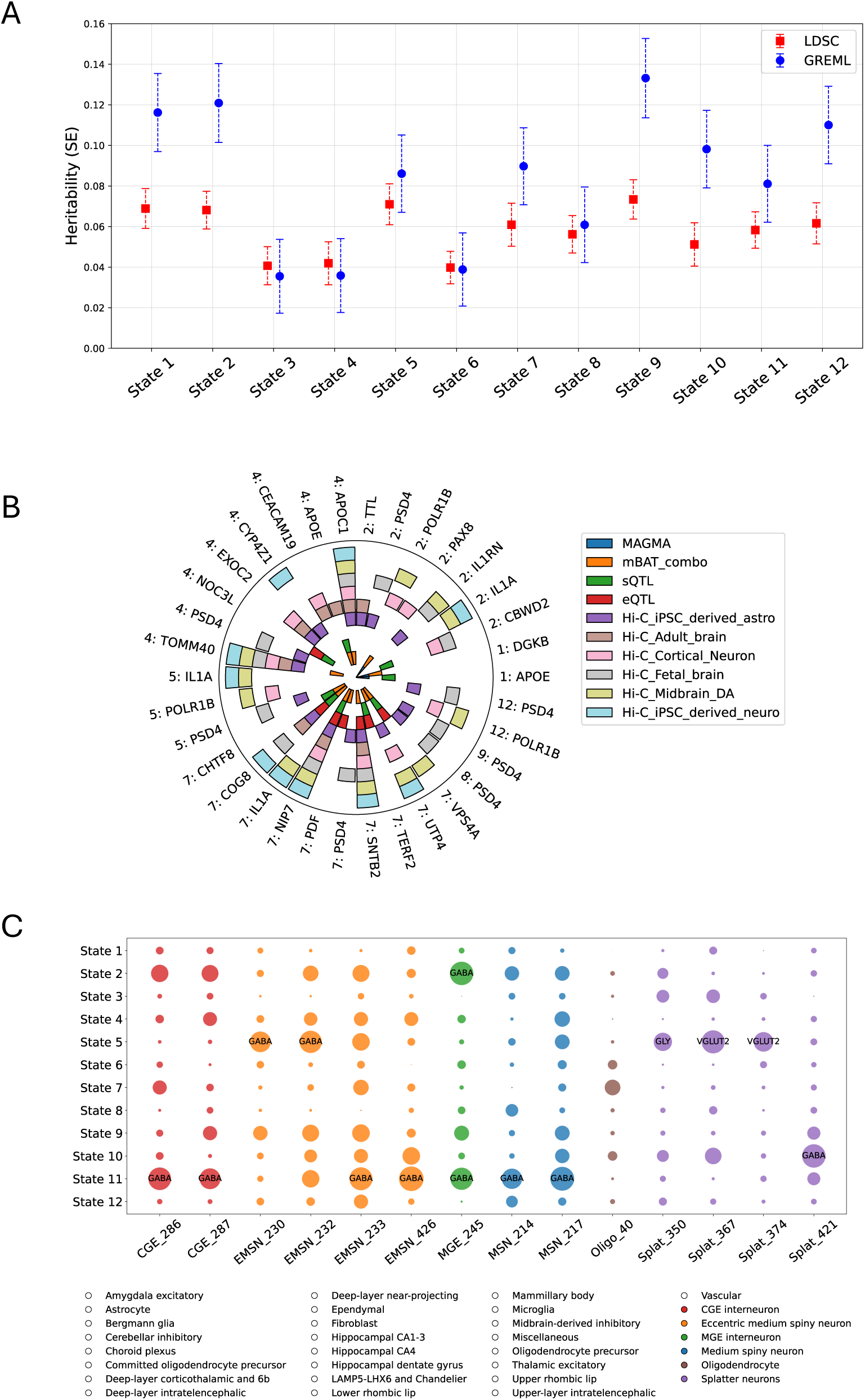
(A) Heritability estimates of brain states derived using two methods: LDSC and GREML. (B) Gene mapping of GWAS results using multiple approaches; shown are genes identified by at least two converging methods. (C) Gene-set enrichment analysis performed with MAGMA, integrating single-nucleus RNA-sequencing data from 461 cell types grouped into 31 superclusters (Siletti et al., 2023). The circle radius represents standardised enrichment scores, and neurotransmitter annotation is added for FDR-corrected ones.

Several complementary genemapping methods were leveraged, including MAGMA [32], H-MAGMA [33], mBAT-combo [34], and QTL mapping [35] (Supplementary Tables 5-8), with loci assigned to genes when at least two approaches converged (Fig. 2B and Supplementary Table 9). We applied functionally informed fine-mapping using PolyFun-SuSiE [36, 37] (*p* < 5 × 10⁻⁸), yet did not limit the QTL mapping to causal SNPs (Supplementary Table 10). In total, 23 genes were identified after FDR-correction, with some states yielding more genes (e.g., 10 in State 7), whereas States 3, 6, 10, and 11 yielded none. The phenotypic relevance of these genes—defined as prior evidence of association with specific traits/diseases—was assessed by querying the GWAS Catalogue. Among overlapping results across states, ***PSD4***—associated with blood cell counts and C-reactive protein (CRP)—emerged in States 2, 4, 5, 7, 8, 9, and 12. ***IL1A***, a cytokine in the interleukin-1 family, appeared in States 2, 5, and 7. ***POLR1B***—linked to CRP levels and smoking-related traits—was identified in States 2, 5, and 12. ***APOE***, widely implicated in neurodegeneration and cognitive performance, was detected in States 1 and 4. All GWAS Catalogue evidence of gene-phenotype associations is provided in Supplementary Table 11.

To gain cellular-level insight into the genetic architecture of brain states, we performed gene-set enrichment analysis with MAGMA using single-nucleus RNA sequencing data comprising 461 cell types grouped into 31 superclusters, together with neurotransmitter annotations for each cell type where available [38] (Fig. 2C and Supplementary Table 12). Across all significant results, enriched cell types included medial and caudal ganglionic eminence interneurons (MGE and CGE), medium and eccentric medium spiny neurons (MSN and EMSN), splatter neurons, and oligodendrocytes. State 2 was enriched for GABAergic MGE interneurons (β = 17.76, *P_FDR_* = 0.045). State 5 showed enrichment across multiple neuronal populations, including GABAergic EMSN and glutamatergic or glycinergic splatter neurons (β range = 10.88–17.23; all *P_FDR_* < 0.05). State 10 was enriched for GABAergic splatter neurons (β = 17.71, *P_FDR_* = 0.0038). State 11 showed enrichment across MSN, EMSN, and both MGE and CGE interneurons, all of which were annotated as GABAergic (β range = 13.37–19.43; all *P_FDR_* < 0.05). Finally, State 7 mapped to oligodendrocytes (β = 8.02, *P_FDR_* = 0.032). This finding supports growing evidence that oligodendrocytes influence cortical processing through metabolic support and that their adaptive myelin plasticity contributes to network synchronisation and dynamics [39, 40]. Overall, these results indicated that dynamic brain states are predominantly mediated by excitatory and inhibitory cell types, with a contribution from oligodendrocytes.

### Brain States Share Polygenic and Pleiotropic Architecture

To address our second aim, we first examined phenotypic and genetic correlations [31] between the FO of brain states (Supplementary Tables 13 and 14). Both correlation matrices showed a similar hierarchical organisation, constructed using the average linkage method (Fig. 3A). The most prominent phenotypic correlation was between States 9 and 2 (*r_p_* = 0.77, SE = 0.0017). Similarly, at the genetic level, States 9 and 2 showed a high correlation (*r_g_* = 0.90, SE = 0.03). Moreover, we found that state-to-state phenotypic correlations were strongly associated with their corresponding genetic correlations (r = 0.90, p = 1.2 × 10⁻²⁴), indicating that phenotypic similarities in FO across brain states are partly driven by shared genetic influences (Fig. 3B).

**Fig. 3.**
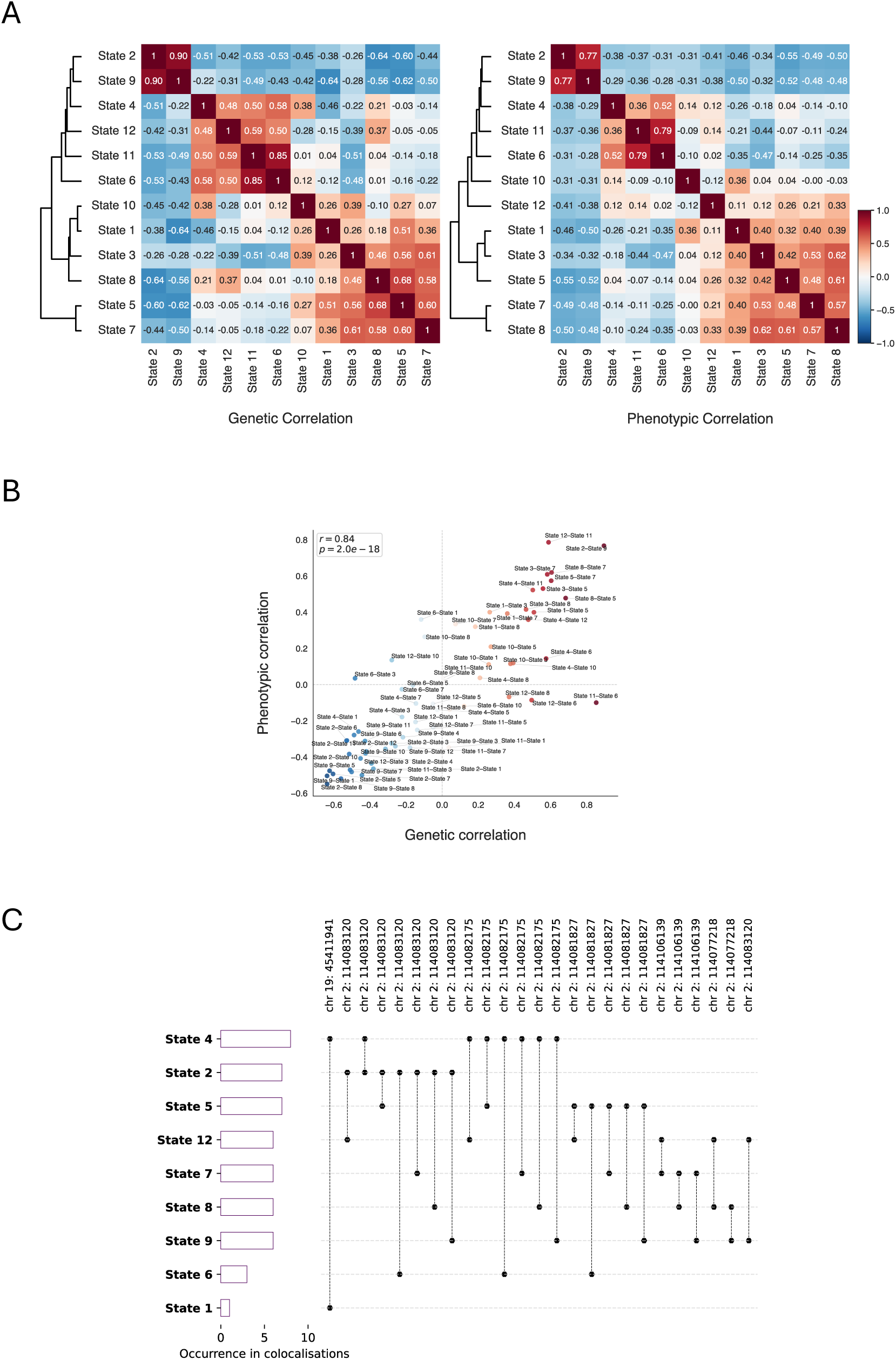
(A) Phenotypic and genetic correlations (estimated using LDSC) between the fractional occupancy (FO) of 12 dynamic brain states. (B) Correspondence between phenotypic and genetic correlation matrices. (C) Between-state genomic colocalisation results using the approximate Bayes factor method; only colocalisations with a posterior inclusion probability (PIP) > 0.95 are shown.

To further characterise the extent to which these shared genetic influences arise from common causal variants rather than genome-wide correlation alone, we performed an approximate Bayes factor colocalisation analysis [41]. All pairwise combinations among the 44 loci were tested, yielding a total of 946 state–state pairs (Supplementary Table 15). Two loci showed particularly strong pleiotropic signals (Fig. 3C). A region on chromosome 2 (113921856–116772470 Mb) near the *PAX8* gene showed widespread colocalisation across States 2, 4, 5, 6, 7, 8, 9, and 12, with PP.H4.abf > 0.97. In addition, a locus on chromosome 19 (44744108-46102697 Mb) within the *APOE* gene cluster demonstrated colocalisation between States 1 and 4 (PP.H4.abf > 0.99). Locus zoom plots are shown (Extended Data Figs. 4 and 5). Taken as a whole, these findings indicate that, despite distinct activation and connectivity profiles, brain states share not only diffuse genome-wide correlations, but also common causal variants.

### Brain States Share Polygenic, Pleiotropic, and Causal Relationships with Broad Phenotypes

Turning to our third aim, we assessed genetic correlations between the FO of brain states and phenotypes with established links to brain function and dysfunction. In light of the view that large-scale brain dynamics reflect the integrated interplay among cognitive, behavioural, and physiological processes, we examined phenotypes across psychiatric, neurological, cognitive, personality, and neuroimaging domains (Fig. 4A and Supplementary Table 16). We observed widespread associations across 576 genetic correlations (12 brain states × 48 phenotypes). Neurological conditions showed relatively large genetic correlations with brain states, particularly dementia with Lewy bodies and vascular dementia, with rg values ranging from −0.37 to 0.43, although none survived FDR correction. Educational attainment had the most associations, involving 10 of 12 brain states (|*r_g_*| up to 0.34; seven survived FDR correction), whereas the personality trait extraversion was correlated with eight states (|*r_g_*| up to 0.30; five survived FDR correction). Sleep duration, schizophrenia, and anxiety each exhibited genetic correlations with five states, although only three or fewer survived FDR correction for each trait.

**Fig. 4.**
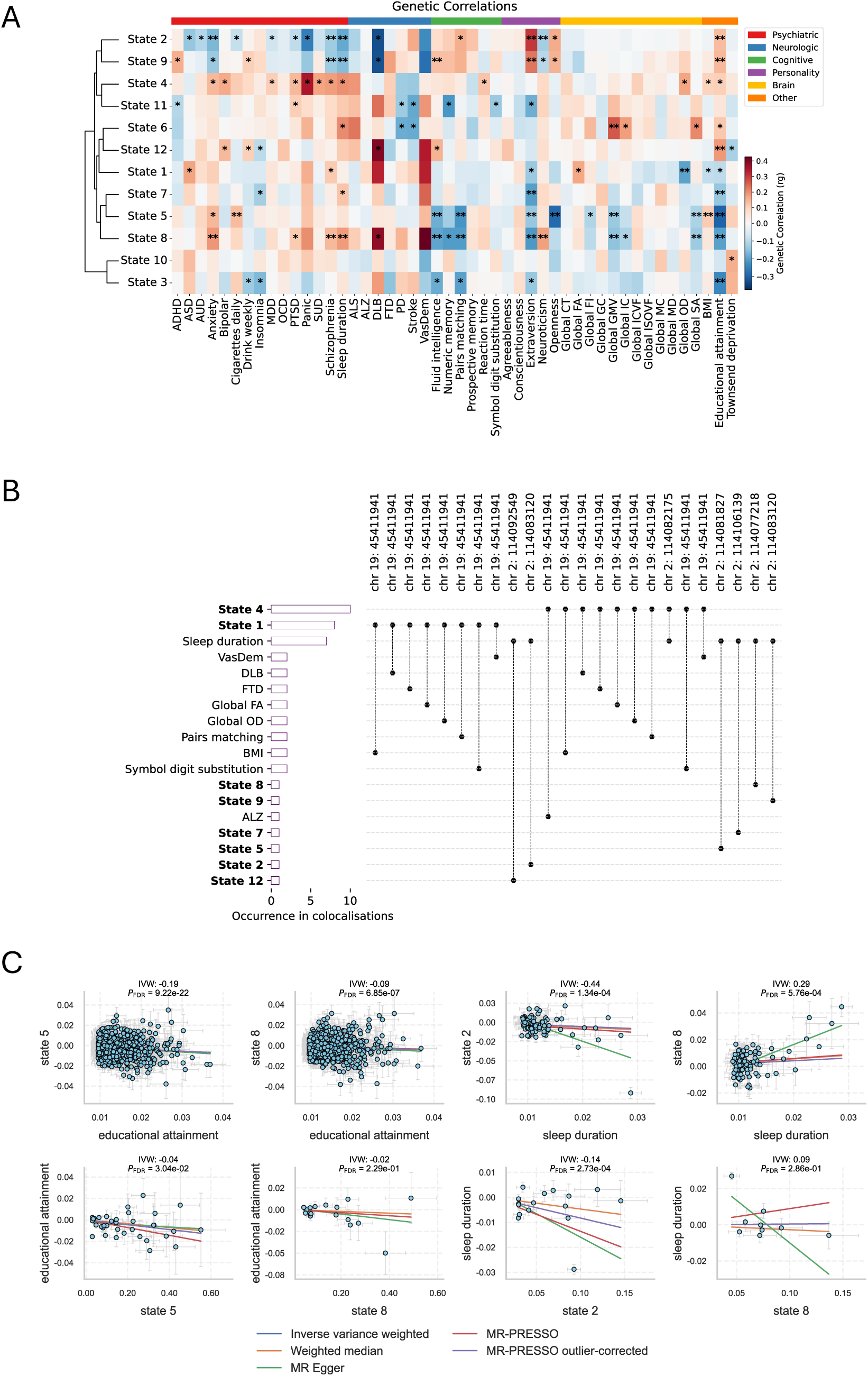
(A) Genetic correlations between the fractional occupancy (FO) of brain states and different phenotype categories. (B) Genomic colocalisation results using the approximate Bayes factor method; only colocalisations with a posterior inclusion probability (PIP) > 0.95 are shown. (C) Forward and reverse Mendelian randomisation analyses; only results meeting the criteria specified in the paper are presented.

To further characterise shared genetic mechanisms, we performed genomic colocalisation analysis across 44 loci and 48 phenotypes, comprising 2,112 pairwise comparisons (Supplementary Table 17). A pleiotropic locus on chromosome 2 (113921856–116772470 Mb) near the *PAX8* showed colocalisation between multiple brain states (2, 4, 5, 7, 8, 9, and 12) and sleep duration, with PP.H4.abf > 0.98 (Fig. 4B). In addition, the pleiotropic locus on chromosome 19 (44744108-46102697 Mb) within the *APOE* gene cluster showed colocalisation between states 1 and 4 and neurodegenerative diseases (Alzheimer’s disease, vascular dementia, dementia with Lewy bodies and frontotemporal dementia), cognitive performance (symbol digit substitution and pairs matching), diffusion MRI measures (fractional anisotropy and orientation dispersion index) and body mass index, with PP.H4.abf > 0.96. Locus zoom plots are shown (Extended Data Figs. 6 and 7). Together, these results demonstrate that shared genetic influences between brain states and complex behavioural and clinical traits are concentrated within biologically meaningful loci.

Finally, Mendelian randomisation (MR) was used to investigate bidirectional causal relationships (Supplementary Table 18). Evidence for causality was accepted when (i) the genetic correlation was significant at P_FDR_ < 0.05, (ii) inverse-variance-weighted (IVW) [42], weighted median [43], MR-Egger [44], and MR-PRESSO [45] estimates were all significant at P_FDR_ < 0.05, (iii) the corrected MR-PRESSO estimate was significant in the presence of horizontal pleiotropy, and (iv) all effect directions were consistent across methods. Across 576 forward and 576 reverse tests, MR analyses indicated that educational attainment and sleep duration exert causal effects on specific brain states (Fig. 4C). Reverse tests met some, but not all, criteria, possibly due to the limited statistical power of the instruments used in the MR for the FO GWAS. We repeated the analyses presented in Fig. 4C using MRlap, which accounts for weak-instrument bias, winner’s curse, and potential sample overlap [46], because UK Biobank participants contributed to both the exposures and outcomes. The MRlap results replicated the original findings, with near-identical P-values (Supplementary Table 19), indicating that the inferred causal effects are robust and unlikely to be driven by sample overlap or weak-instrument bias. We further addressed the robustness of findings. MR-Egger intercepts showed no directional pleiotropy for educational attainment (all P > 0.05), but significant pleiotropy for sleep duration (Supplementary Table 20). Cochran’s Q tests for both IVW and MR-Egger indicated significant heterogeneity across instruments for all the associations above. These results suggest biologically heterogeneous genetic effects, meaning MR estimates should be interpreted as average causal effects rather than a single mechanism. Notably, consistent effect directions and significance across multiple MR methods support the robustness of the inferred causal relationships despite heterogeneity and pleiotropy.

### Cross-ancestry replication

We used non-European UKB sub-samples for cross-ancestry validation. We assessed the sign concordance of clumped lead SNPs between the European discovery sample and non-European populations, as well as the correlation between their beta estimates (Supplementary Table 21 and Extended Data Fig. 8), including only SNPs with a minor allele frequency > 0.001 in the replication population. Among variants shared between European and East Asian (EAS; n = 189) ancestries, 10 of 12 lead SNPs (83.3%) had concordant effect directions, with beta estimates showing a positive Spearman correlation (r = 0.51). In comparisons between European and South Asian (SAS; n = 728) ancestries, 12 of 21 lead SNPs (57.1%) were concordant, with beta estimates again showing a positive correlation (r = 0.58). Despite limited power due to the small sample sizes of the replication groups, these results indicate partial replication of genetic effects across ancestries. We did not perform a trans-ancestry meta-analysis because of markedly imbalanced sample sizes across ancestry groups and differences in allele frequencies and linkage disequilibrium patterns, which could bias meta-analytic estimates.

## Discussion

The study investigated the genetic architecture of dynamic brain states derived from resting-state fMRI using a Hidden Markov Model (HMM) framework. By combining large-scale neuroimaging data from over fifty thousand individuals with genome-wide association analyses, we aimed to determine whether the temporal properties of brain states are genetically influenced, whether distinct states share common genetic mechanisms, and whether their genetic architecture overlaps with that of traits and diseases.

The primary objective of this study was to determine whether common genetic variants can predict dynamic brain states. SNP-based estimates indicate that a non-trivial proportion of inter-individual variability in fractional occupancy (FO) is attributable to common genetic variation, with effect sizes comparable to those reported for other imaging-derived phenotypes (IDPs) [12–20]. The heritability results thereby position dynamic brain states alongside traditional IDPs as genetically informative traits. Beyond heritability, the study identifies 44 independent loci and 23 genes associated with FO across states. Several genes—such as ***APOE***, ***APOC1, DGKB, PAX8, CBWD2 and NOC3L***—have been repeatedly implicated in neuroimaging [12–20] or neurological phenotypes [47, 48], reinforcing the validity of dynamic states as meaningful neural constructs.

Cell-type enrichment analyses provided insight into the neurobiological substrates underlying these genetic associations. Across states, significant enrichment was observed in several neuronal populations, particularly GABAergic interneurons and medium spiny neurons, as well as in oligodendrocytes. GABAergic interneurons play a key role in regulating cortical excitability [49], while medium spiny neurons, the principal cells of the striatum, integrate cortical input and modulate large-scale network activity through basal ganglia–thalamocortical circuits [50, 51]. In addition, enrichment in oligodendrocytes supports the idea of myelin plasticity in coordinating large-scale neural communication [39, 40]. Together, these findings indicate that dynamic brain states are supported by interactions among multiple cellular systems, including inhibitory signalling, subcortical circuits, and white matter processes.

As a second aim, we demonstrated that brain states exhibit polygenic and pleiotropic genetic architecture. Phenotypic correlations in FOs were strongly mirrored by genetic correlations, showing similar patterns in how states cluster. Colocalisation analyses further revealed several loci, most notably regions near PAX8 and APOE, that harbour shared causal variants influencing multiple states. Of note, while the concept of functional hierarchy has been explored previously [52, 53], our study advances the field in two ways. First, prior analyses typically assume a hierarchy a priori, whereas the HMM neither imposes nor encourages one, meaning that any hierarchy to emerge naturally from the data. Second, we showed that the hierarchy of states is not only evident at the phenotypic level but is also grounded in genetic architecture.

The third aim was to investigate the genetic overlap between the FO of brain states and a set of phenotypes relevant to normal or pathological brain function. Several brain states were found to be genetically correlated with educational attainment and sleep duration, while Mendelian randomisation analyses indicated causal effects on brain dynamics. These observations accord with prior work linking educational attainment to brain structure [54], functional connectivity [55], and cognitive reserve [56]. Sleep, in turn, has been shown to modulate synaptic plasticity [57], metabolic clearance [58], and the organisation of large-scale brain networks [59].

Interestingly, the associations between state-specific FOs and the investigated phenotypes may arise from shared biological pathways, as supported by convergent genetic signals. We found a locus near ***PAX8*** that was observed across sleep duration and multiple brain states. Considering the role of *PAX8* in thyroid regulation [59], endocrine mechanisms may jointly influence sleep regulation and the temporal organisation of brain states. Another interesting finding was the ***APOE*** locus, which revealed shared association signals between brain states and several neurodegenerative diseases, including Alzheimer’s disease, vascular dementia, and dementia with Lewy bodies. Given the well-established roles of **APOE** in lipid metabolism [60], synaptic function [61], and neurodegeneration [62], dynamic brain states may serve as functional intermediates that link molecular processes to the large-scale network disruptions observed in neurodegenerative disorders. Longitudinal studies will be essential to determine whether changes in FO precede clinical onset or track disease progression.

Last but not least, our cross-ancestry analysis provided encouraging, though modest, evidence of generalisability. Despite small sample sizes, the direction of effects was largely concordant across European, East Asian, and South Asian groups. This suggests that at least some genetic influences on dynamic brain states are shared across populations, though larger and more balanced multi-ancestry datasets will be essential for robust inference.

## Method

### Genotype

We used imputed genotype array data from the UK Biobank. A detailed description of genetic pre-processing is provided elsewhere [21, 22]. Analyses were restricted to participants of self-reported European ancestry. Subjects were excluded if they exceeded ±5 standard deviations on the first two genetic principal components, had a genotyping rate <95%, displayed excessive heterozygosity, or showed discordance between genetic and self-reported sex. Variants were filtered to retain those with minor allele frequency >1%, call rate >95%, Hardy–Weinberg equilibrium p > 1 × 10⁻⁶, and, for imputed SNPs, imputation r² > 0.4. Following quality control, 19,914,312 SNPs were included in the final analysis. Other ancestry groups were excluded due to small sample sizes and the absence of complete genetic and phenotypic data, which together resulted in insufficient statistical power for mixed-effects genome-wide association analyses. After integrating genetic and imaging data, 52,335 individuals were included in the final analysis.

### Imaging

UK Biobank resting-state fMRI data were used, acquired on a 3T Siemens Skyra scanner (multiband = 8; 490 time points; TR = 0.735 s; 2.4 mm isotropic resolution). The standard UK Biobank preprocessing pipeline included motion correction, EPI distortion correction, temporal filtering, and FMRIB’s ICA-based X-noisefier (FIX) [63]. We applied imaging quality control prior to analysis, and participants were excluded if mean framewise displacement (FD) [64], maximum FD, or Euler index [65] exceeded five mean or median absolute deviations from the sample distribution.

Group ICA results obtained using FSL’s MELODIC were employed, with 100 components initially estimated. The resulting ICA spatial maps were treated as functional parcellations of cortical and subcortical grey matter. Components clearly identifiable as artefactual (i.e., not neuronally driven) were discarded, yielding 55 non-artefactual components. The retained ICA spatial maps were projected onto subject-specific fMRI data using dual regression analysis to derive a representative time series per component, which were then used in subsequent analyses.

We used Hidden Markov Modelling (HMM) to analyse resting-state fMRI data [5, 6]. Let x_t_ denote the observed data and s_t_ the hidden state at time point *t*. This can be modelled as:

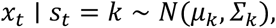

where μ_k_ is a vector of mean BOLD activations across channels (e.g., brain areas), and Σ_k_ is the covariance matrix capturing both variances and covariances for state *k*. This specification is known as the observation model, as it defines the probabilistic distribution of the data associated with each state through the parameters (μ_k_, Σ_k_). It should be noted that alternative observation models are possible; for example, a multivariate autoregressive (MAR) model, in which states are distinguished by their spectral signatures [48]. The temporal sequence of states is further constrained by a transition model that encodes the probability of moving from one state to another. Specifically, the probability that state *k* is active at time *t* depends on the state at the previous time point, *t − 1*, according to:

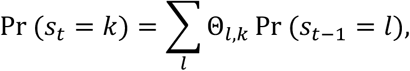

where Θ_l,k_ represents the transition probability from state *l* to state *k*. The diagonal elements of Θ (Θ_kk_) control state persistence, while off-diagonal elements (Θ_k__l_, with k ≠ l) represent transitions between distinct states. Additionally, the parameter η defines the probability of each state at the start of a session. Consequently, the observed data at any time point can be interpreted as a mixture of Gaussian distributions, with the mixture weights given by w_t__k_ = Pr (s_t_ = k).

Estimating the observation model in the Gaussian case requires inverting a Q-by-Q matrix per state, with Q being the number of time series. Standard variational inference demands the full dataset in memory and computes state time courses via Baum-Welch recursions [66], which is costly for large datasets. To address this, we employed stochastic variational inference, updating parameters using subsets of M subjects, computing interim state observation models, and combining them with the current estimates. This update follows (1):

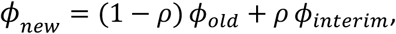

where Φ*_new_*, Φ*old*, and Φ*_interim_*are the posterior distributions of the observation models at the current, previous, and interim updates, respectively, and ρ decreases over iterations according to (2):

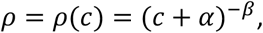

with *c* the current iteration, along with α and β representing «delay» and «forget» parameters. State transition probabilities are updated exactly using Σ*_t_ Pr Pr* (*S_t_*) *Pr* (*S_t-1_*), where *S_t_* is the hidden state at time *t*, and *Pr* (*S_t_*) its probability. Subject selection is biased toward those less frequently sampled using (3):

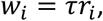

where *w_i_* is the unnormalised probability of selecting subject *i*, *r_i_* the number of times subject *i* has been previously selected (scaled so that *min* (*r_i_*) = 0), and τ ≤ 1 controls the degree of discouragement.

To improve computational efficiency and avoid poor local minima, an initialisation strategy was used, running standard HMM inference on subsets of subjects and merging the results. The algorithm iteratively selects M subjects based on *w_i_*, computes their state time courses, estimates interim state observation models as if N subjects were used, updates approximate posterior distributions via (1), updates exact state transitions, and adjusts ρ via (2) until free energy convergence. Five inference runs with random initialisations were performed, and the solution with the highest free energy was used for the final model.

### Network-based spatial co-location

To clarify the spatial configuration of brain states, we mapped them onto seven canonical functional networks [23]: visual, somatomotor, dorsal attention, ventral attention, limbic, frontoparietal, and default mode. For this purpose, group-level activation maps were parcellated using the Glasser atlas, which provides a high-resolution, multimodal cortical parcellation dividing each hemisphere into 180 regions [24]. Parcellation and extraction of regional mean values were performed using the Neuromaps [67]. For each brain state, we computed Pearson correlations between the mean activation of Glasser parcels and their corresponding canonical network assignments. To account for the inherent spatial autocorrelation in cortical data—which can artificially inflate correlation estimates due to neighbouring regions being spatially smooth—we employed a spin permutation procedure [25]. In this approach, the cortical parcellation is projected onto a spherical surface and randomly spun 10,000 times to generate a null distribution of correlations that preserves the spatial covariance structure of the cortex. Empirical correlations were then compared against this null distribution to determine statistical significance.

### Genome-wide association study (GWAS)

The fastGWA model [27] is a linear mixed model that can be expressed as:

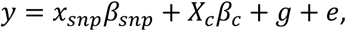

where *y* is a vector of mean-centred phenotypes, *x_snp_* represents the mean-centred genotypes for the variant being tested with effect β_snp_, *X*_c_ is the matrix of fixed covariates such as age, sex, and ancestral diversity with coefficients β_c_, *g* denotes the polygenic effect captured through familial relatedness and follows a normal distribution *g ∼ N(0, πσ^2^_g_)*, and *e* is the residual error term distributed as *e ∼ N(0, πσ^2^_e_)*. The matrix π encodes family relatedness based on pedigree information, and the overall variance-covariance matrix of the phenotypes is given by *V = πσ^2^_g_ + Iσ^2^_e_*. When pedigree information is missing or incomplete, π can be replaced by a sparse SNP-derived genetic relationship matrix (GRM), i.e., small off-diagonal elements below a threshold (e.g., 0.05) set to zero. The variance components σ^2^_g_ and σ^2^_e_ are unknown and are estimated using restricted maximum likelihood (REML). fastGWA implements a grid-search REML algorithm, called fastGWA-REML, which avoids the computationally intensive inversion of the variance-covariance matrix. Once the variance components are estimated, the SNP effect can be calculated using generalised least squares:

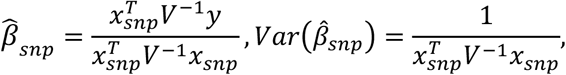

Notably, the procedure achieves substantial computational efficiency gains by employing the GRAMMAR-GAMMA approximation. Benchmarking on UK Biobank data has shown that fastGWA is faster and far more memory-efficient than existing mixed-model approaches, such as BOLT-LMM.

To account for relatedness and population structure, we computed a sparse genetic relatedness matrix (GRM) from LD-pruned SNPs and derived genetic principal components (PCs) using fastPCA [28] implemented in the SNPRelate package [68]. All phenotypes were converted to *z* scores to ensure comparability. We then conducted 12 GWAS, one for each phenotype, adjusting for covariates including the first 16 genetic PCs, age, age², sex, age×sex, age²×sex, imaging site, mean and maximum framewise displacement, and the Euler index. Independent signals were identified using PLINK clumping with a genome-wide significance threshold of p < 5 × 10⁻⁸, a 1-Mb physical distance window, and an LD cutoff of r² = 0.1. Replication and cross-ancestry generalisability were assessed by comparing the direction of effects for clumped lead SNPs from the European discovery cohort with those observed in East Asian (EAS; n = 189) and South Asian (SAS; n = 728) ancestry groups, and by assessing correlations between effect-size estimates.

### Heritability

Heritability for each GWAS of brain states was estimated using both LD Score Regression (LDSC) [31] and Genome Restricted Maximum Likelihood (GREML) [30]. LDSC relies on GWAS summary statistics and does not require individual-level genotype data. Under a polygenic model, SNPs in LD with causal variants are expected to show inflated test statistics proportional to their LD, whereas inflation arising from confounding factors such as cryptic relatedness or population stratification is independent of LD. By regressing the χ² statistic of each SNP on its LD score—the sum of squared correlations (r²) with surrounding variants—the slope estimates SNP-based heritability, while the intercept minus one quantifies the contribution of confounding. This framework enables effective separation of true polygenic signal from systematic bias. We used European reference LD scores derived from the 1000 Genomes Project [69].

In the GCTA-GREML framework, each individual’s phenotype is modelled as a function of additive genetic effects and residual error, with the total genetic effect defined as the sum of effects across causal loci. Because the true causal variants are generally unknown, a genetic relationship matrix (GRM) is constructed to proxy for the unobserved relationships at causal loci. Accuracy improves by adjusting the GRM with a weighted regression approach that accounts for sampling error and minor allele frequency. Restricted maximum likelihood (REML) is applied to the adjusted GRM to estimate the additive genetic variance. This procedure quantifies the proportion of phenotypic variance attributable to genetic effects.

### Gene mapping

We applied multiple gene-mapping approaches to link GWAS signals to their target genes. First, we used MAGMA [32], which aggregates SNP-level associations into gene-level statistics while accounting for confounders. Despite its widespread use, MAGMA has notable limitations, as it assigns SNPs to the nearest genes and therefore overlooks long-range regulatory interactions through which non-coding variants can influence distal genes. In addition, MAGMA does not incorporate tissue-specific regulatory relationships, even though disease-associated variants are usually enriched in regulatory elements of relevant tissues.

To overcome the limitations of MAGMA, we also employed H-MAGMA [33], which integrates chromatin interactions to improve SNP-to-gene mapping. In H-MAGMA, exonic and promoter SNPs are assigned based on genomic proximity, whereas intronic and intergenic SNPs are linked to genes through experimentally derived chromatin interactions. We used Hi-C data from foetal brain [70], adult brain [71] and two cell-type–specific datasets derived from midbrain dopaminergic [72] and cortical neurons [73], enabling us to capture regulatory interactions that are both developmentally informed and cell-type specific.

As a third approach, we applied a gene-based testing strategy to improve the detection of multi-SNP associations. Conventional gene-based tests typically combine SNP associations by summing χ² statistics, an approach that ignores SNP effect direction and can therefore lose power in the presence of masking, such as when SNP effect sizes and their LD correlations act in opposite directions. To overcome this limitation, we used mBAT-combo [34], which does not rely on directional consistency but instead tests whether a set of SNPs accounts for a nonzero proportion of phenotypic variance. By adopting this variance-based framework, mBAT-combo offers greater statistical power than traditional gene-based methods, particularly in the context of complex LD structures.

As a final approach, we conducted QTL-based mapping by linking genetic variants to their downstream regulatory effects. We queried expression QTLs (eQTLs) and splicing QTLs (sQTLs) derived from RNA sequencing data of 2,865 human brain cortex samples aggregated across seven cohorts [74]. In addition, we incorporated DNA methylation QTLs (mQTLs) from a previous meta-analysis (estimated effective n = 1160) [75], which integrates data from three cohorts. We also included chromatin accessibility QTLs (caQTLs) derived from prefrontal cortex samples from 135 patients with schizophrenia and 137 controls [76]. Together, these QTL resources enabled a comprehensive assessment of how GWAS variants influence gene expression, splicing, epigenetic modification, and chromatin accessibility in the human brain.

### Fine-mapping

We applied PolyFun + SuSiE to all genome-wide significant variants (p < 5 × 10⁻⁸) to identify likely causal SNPs [36]. PolyFun prioritises putative causal variants by integrating prior probabilities into fine-mapping methods such as SuSiE [37] and FINEMAP [77]. Functional enrichments are estimated across a broad set of coding, conserved, regulatory, and LD-related annotations from the baseline-LF model using an L2-regularised extension of stratified LD score regression (S-LDSC) [78]. Per-SNP heritabilities are then inferred in a cross-chromosome framework, excluding the target chromosome to mitigate the winner’s curse and improve robustness to model misspecification. Prior causal probabilities for SNPs within each locus are set proportional to these refined per-SNP heritability estimates. In simulations, PolyFun, when combined with SuSiE or FINEMAP, was well calibrated and identified more than 20% more high-confidence causal variants than non–functionally informed fine-mapping approaches.

### Cell data

We utilised the most extensive human brain single-nucleus RNA sequencing dataset available [38], which includes 3,369,219 nuclei from 105 brain regions, collected from roughly 100 dissections spanning the forebrain, midbrain, and hindbrain of three postmortem donors. These nuclei were organised into 31 superclusters and 461 clusters—considered as distinct cell types—capturing both developmental origins and regional variation. Rather than assigning a single label to each cluster, a multilayered auto-annotation approach was employed. Using the Linnarsson lab’s framework (https://github.com/linnarsson-lab/auto-annotation-ah), clusters were annotated across four categories: neurotransmitters, neuropeptides, broad classes, and more specific subtypes. This comprehensive transcriptomic map of the human brain serves as a valuable resource for exploring molecular diversity in both health and disease.

### Cell-type enrichment

MAGMA was used to translate the genetic basis of brain states into cellular components. SNPs were mapped to genes if located within 35 kb upstream and 10 kb downstream, with the MHC region excluded due to high LD. Gene-level associations were computed using the SNP-wise mean model, with LD adjusted based on the European ancestry panel from the 1000 Genomes Project. Specificity scores were calculated to quantify the fraction of each gene’s total expression in each cell type; values ranged from 0 to 1, and the scores summed to 1 across the 461 cell types for each gene [79]. MAGMA gene property analysis was then applied using multiple linear regression to determine whether genes associated with a brain state were preferentially expressed in specific cell types. Covariates, including gene size, gene density, sample size, and minor allele counts, were included in the regression. Multiple comparisons were adjusted for FDR.

### Genetic correlation

Genetic correlations between traits were estimated using LDSC [31]. For each SNP, the product of Z-scores across the two traits is regressed on the SNP’s LD score, thereby capturing the genome-wide covariance of effect sizes. Confounding factors, including population stratification, are efficiently accounted for through the regression intercept. We used European reference LD Scores derived from the 1000 Genomes Project to ensure compatibility with our GWAS cohort.

### Genomic co-localisation

Colocalisation was assessed using an Approximate Bayes Factor (ABF) framework [41]. For each GWAS pair, the sentinel SNP of the clumped GWAS was used to define the corresponding LD block, from which all SNPs were extracted. Analyses were restricted to SNPs shared within this region across both traits, and colocalisation was then evaluated. In this method, for each SNP, an ABF quantifies the relative support for association with a trait against the null model of no association, using either p-values or regression coefficients and their variances. A “configuration” is a pair of binary vectors indicating which SNPs are causal for each trait, with at most one causal SNP per trait. All possible configurations are grouped into five hypotheses: PP0 (no association with either trait), PP1 and PP2 (association with only one trait), PP3 (two independent causal SNPs), and PP4 (a single shared causal SNP). Posterior probabilities for each hypothesis are computed by summing the ABFs across all configurations consistent with that hypothesis, with default prior probabilities used in our analyses. Large PP4 values indicate colocalisation, while large PP3 values indicate independent associations.

### Mendelian Randomisation

Mendelian randomisation (MR) uses genetic variants as instruments to estimate the causal effects of exposures on outcomes [80]. It rests on three key assumptions: the variants must be strongly associated with the exposure, independent of confounders, and influence the outcome solely through the exposure [81]. Multiple MR estimators and sensitivity analyses are usually applied to test these assumptions and strengthen the validity of causal inference.

We applied Mendelian randomisation (MR) to investigate bidirectional causal relationships between brain states and various phenotypes, conducting 576 forward (12 brain states × 48 phenotypes) and 576 reverse tests. Genetic instruments were selected by clumping at P < 5 × 10⁻⁸, with a lenient threshold (P < 5 × 10⁻⁶) for brain-state exposures given the limited number of genome-wide significant variants. Each exposure–outcome pair was filtered to retain only clumped SNPs and harmonised for allele alignment. The MR-Steiger test was used to assess whether the data supported the proposed directionality [82]. If the test indicated an incorrect direction, Steiger filtering was applied to remove SNPs that explained more variance in the outcome than in the exposure. Analyses proceeded only when at least five SNPs remained after these steps.

Causal effects were then estimated using inverse-variance-weighted (IVW) [42], weighted median [43], MR-Egger [44], and Mendelian Randomisation Pleiotropy Residual Sum and Outlier (MR-PRESSO) [45]. IVW provides primary estimates; the weighted median remains valid if ≥50% of instruments are valid, MR-Egger accounts for directional pleiotropy, and MR-PRESSO corrects for outliers. In our study, causality was supported when significant genetic correlations coincided with significant effects across all MR methods, consistent effect directions, and significant corrected MR-PRESSO estimates where horizontal pleiotropy was detected. For results that satisfied the conditions described above, we further applied MRlap [46], which uses cross-trait LD score regression (LDSC) to estimate sample overlap and produce bias-corrected causal effects. We assessed the reliability of causal estimates using Cochran’s Q for heterogeneity, MR-Egger intercept for directional pleiotropy, and MR-PRESSO global test for SNP outliers.

## Ethics Statement

This research has been conducted using the UK Biobank Resource under application number 20904, and utilised anonymised data under the existing Research Tissue Bank (RTB) approval.

## Data availability

GWAS summary statistics for **12 dynamic brain states** will be made available upon reasonable request to the corresponding author. Additionally, we obtained GWAS summary statistics for a wide range of phenotypes, including **psychiatric disorders/traits** (attention-deficit/hyperactivity disorder (ADHD) [83], alcohol use disorder (AUD) [84], anxiety [85], autism spectrum disorder (ASD) [86], bipolar disorder [87], cigarettes per day [88], drinks per week [88], insomnia [89], major depressive disorder (MDD) [90], obsessive–compulsive disorder (OCD) [91], panic [92], post-traumatic stress disorder (PTSD) [93], schizophrenia [94], sleep duration [95], and substance use disorder (SUD) [96]), **neurologic diseases** (Alzheimer’s disease (ALZ) [97], amyotrophic lateral sclerosis (ALS) [98], dementia with Lewy bodies (DLB) [99], frontotemporal dementia (FTD) [100], Parkinson’s disease (PD) [101], stroke [102], and vascular dementia (VasDem) [103]), **cognitive performance** (fluid intelligence, numeric memory, pairs matching, prospective memory, reaction time, and symbol digit substitution; all from [95]), **personality traits** (agreeableness, conscientiousness, extraversion, neuroticism, and openness; all from [104]), and **others** (body mass index (BMI) [95], educational attainment [105], and the Townsend deprivation index [95]).

For whole-brain imaging phenotypes, we used GWAS summary statistics from an updated version of our previous GWAS [20], which has not yet been published. The global brain imaging phenotypes included **macrostructural metrics**—surface area (SA), gray matter volume (GMV), cortical thickness (CT), folding index (FI), intrinsic curvature index (ICI), mean curvature (MC), and Gaussian curvature (GC)—as well as **microstructural measures**—fractional anisotropy (FA), mean diffusivity (MD), isotropic volume fraction (ISOVF), intracellular volume fraction (ICVF), and orientation dispersion (OD).

## Code availability

The analysis pipeline is publicly available here: (https://github.com/amir-ebneabbasi/Brain-dynamics-GWAS).

---

**Extended Data Fig. 1.**
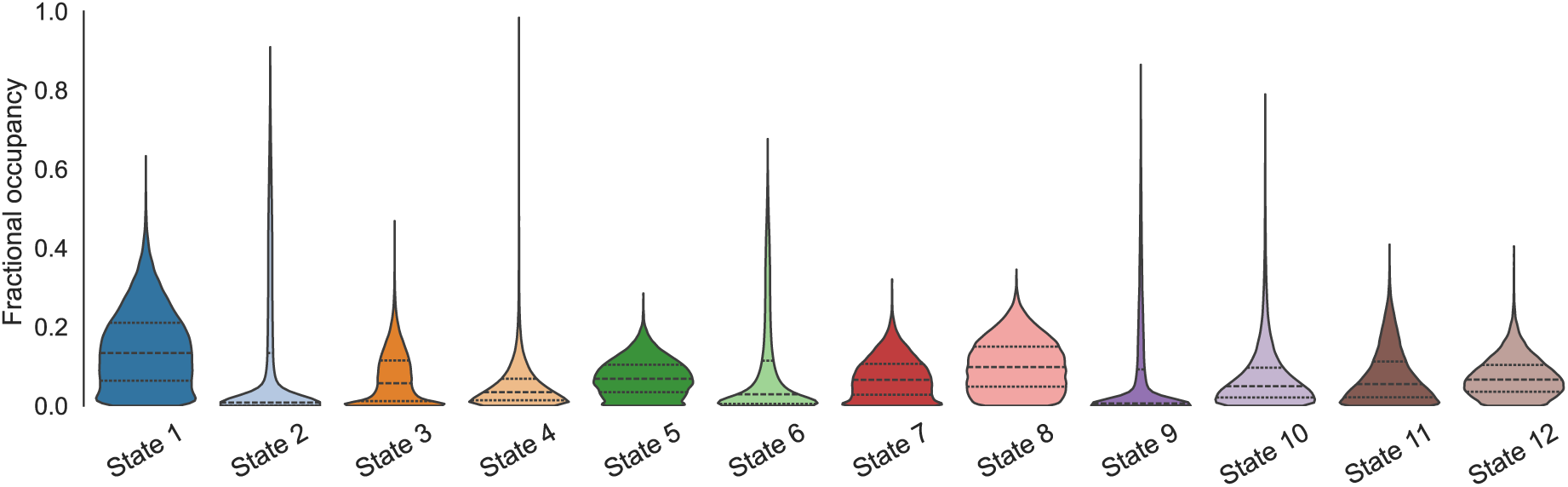
Violin plots showing the distribution of fractional occupancy in the 12 brain states in the UK Biobank cohort. The median (50th percentile), as well as the 25th (Q1) and 75th (Q3) percentiles, are indicated.

**Extended Data Fig. 2.**
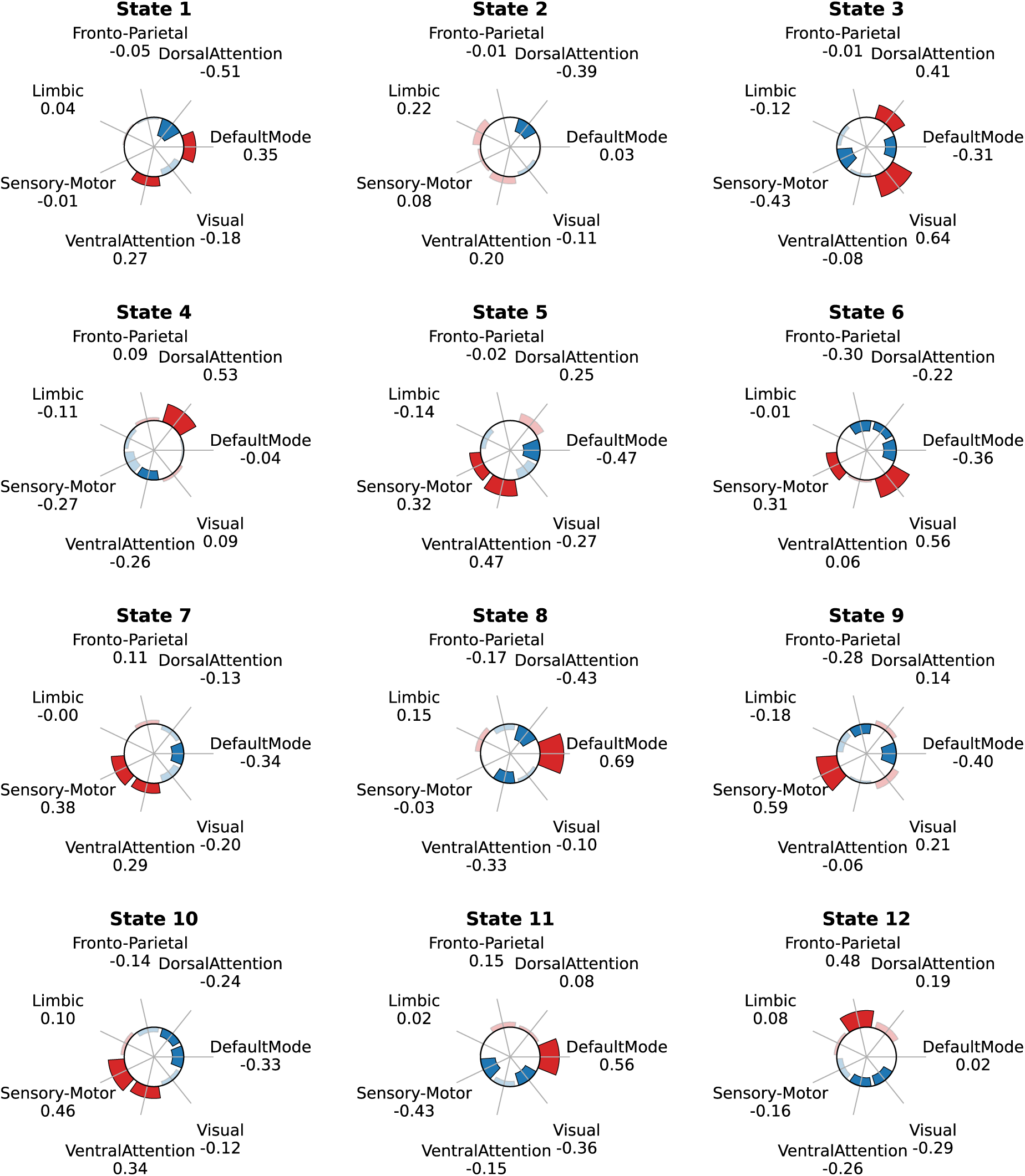
The spatial co-location of HMM-derived brain states with Yeo’s seven networks. Significant associations are shown in brighter colours, with red indicating positive associations and blue indicating negative associations.

**Extended Data Fig. 3.**
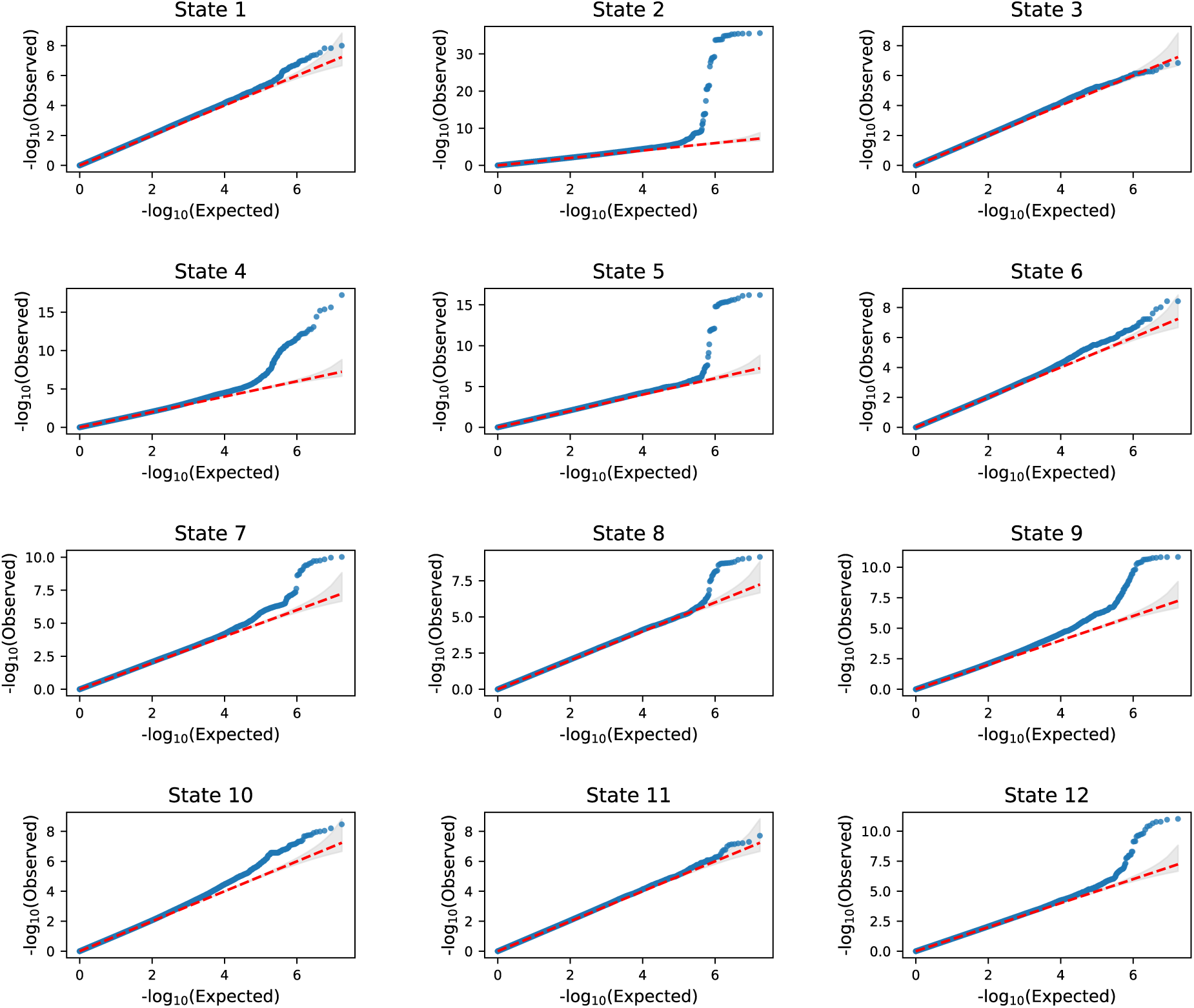
Quantile–quantile (QQ) plots for the GWAS of the 12 brain states, showing the distribution of observed versus expected p-values for each state.

**Extended Data Fig. 4.**
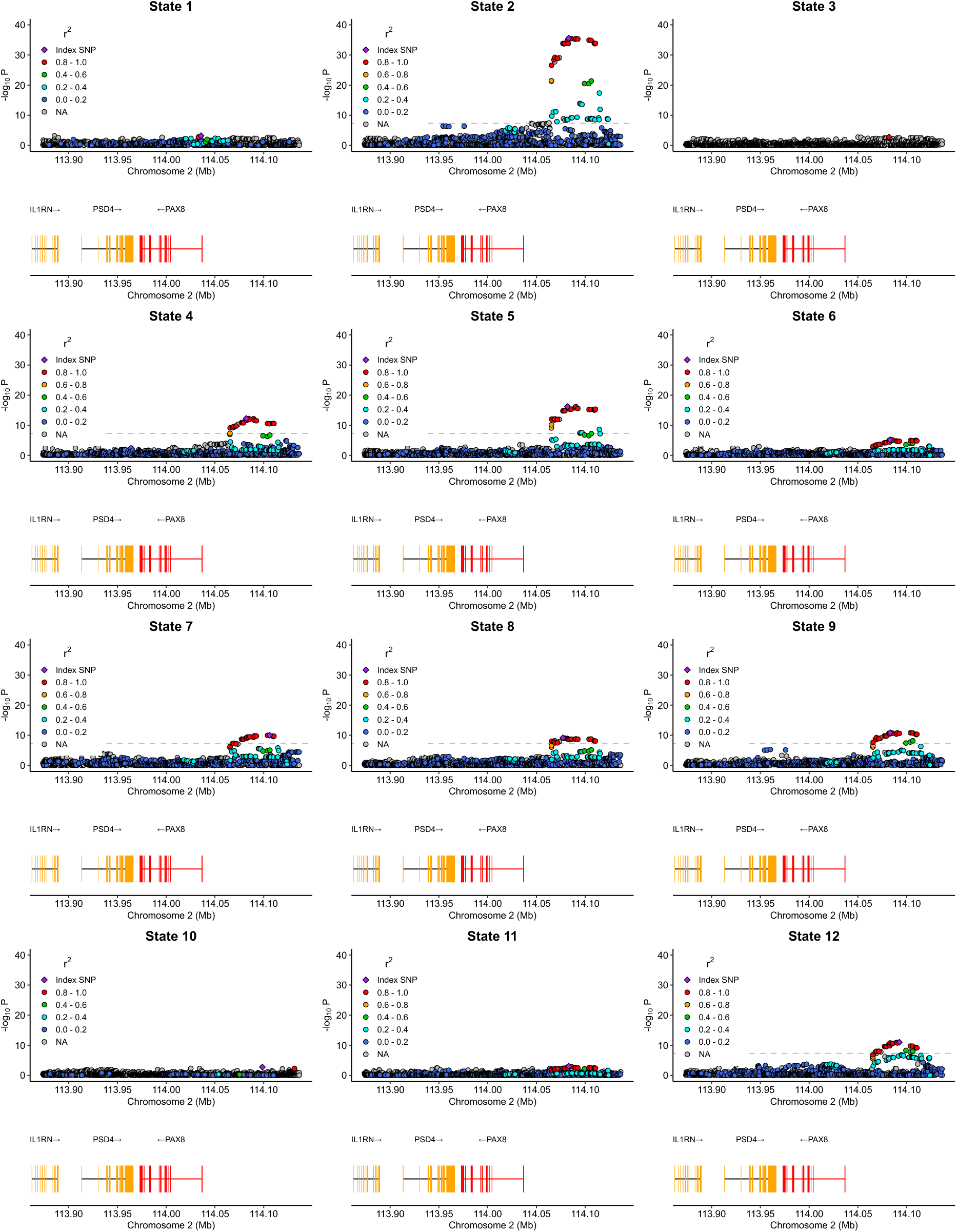
Shared genetic signals near **PAX8** on chromosome 2 across brain states 2, 4, 5, 6, 7, 8, 9, and 12 (PP.H4.abf > 0.95).

**Extended Data Fig. 5.**
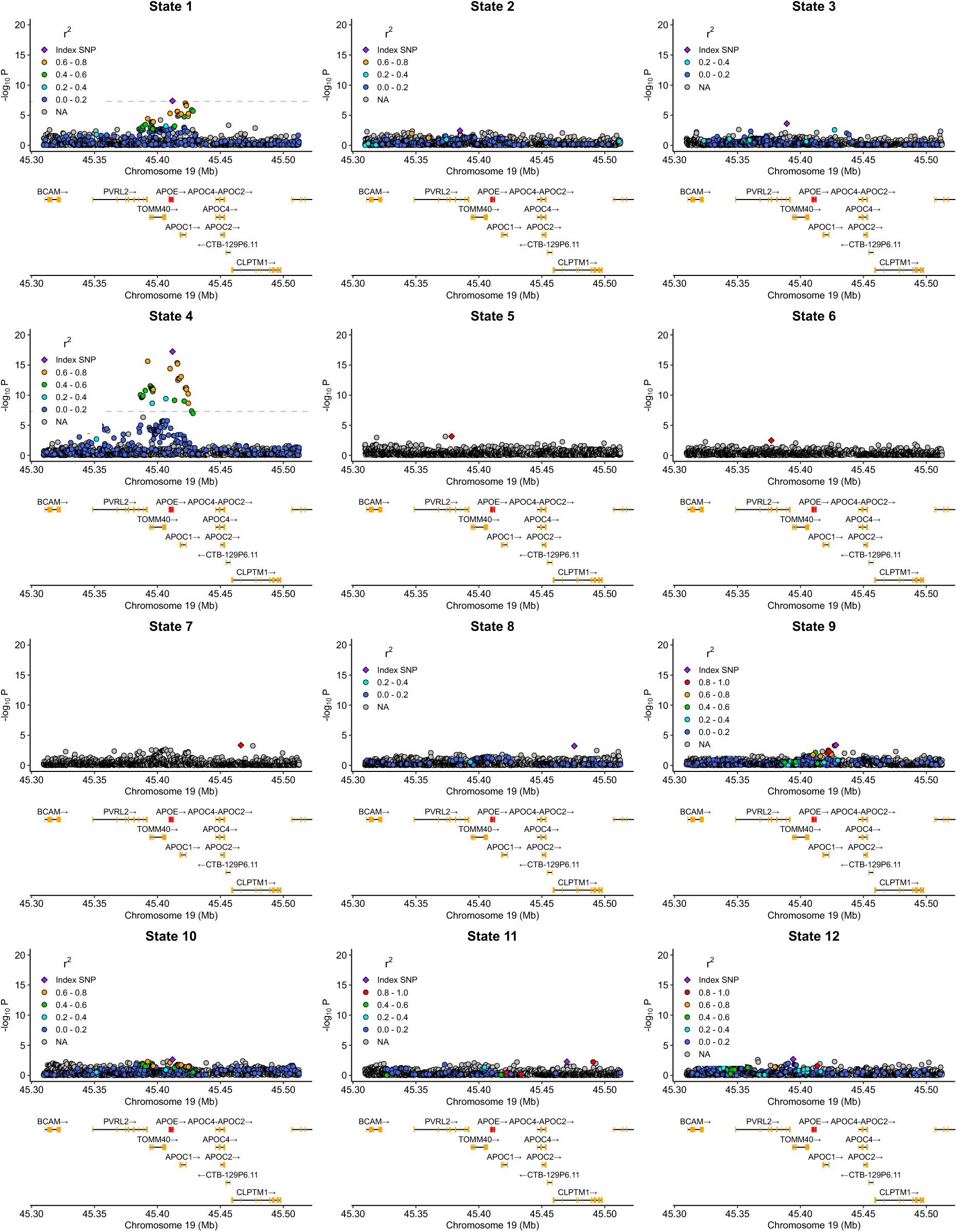
Shared genetic signals within **APOE** on chromosome 19 across brain states 1 and 4 (PP.H4.abf > 0.95).

**Extended Data Fig. 6.**
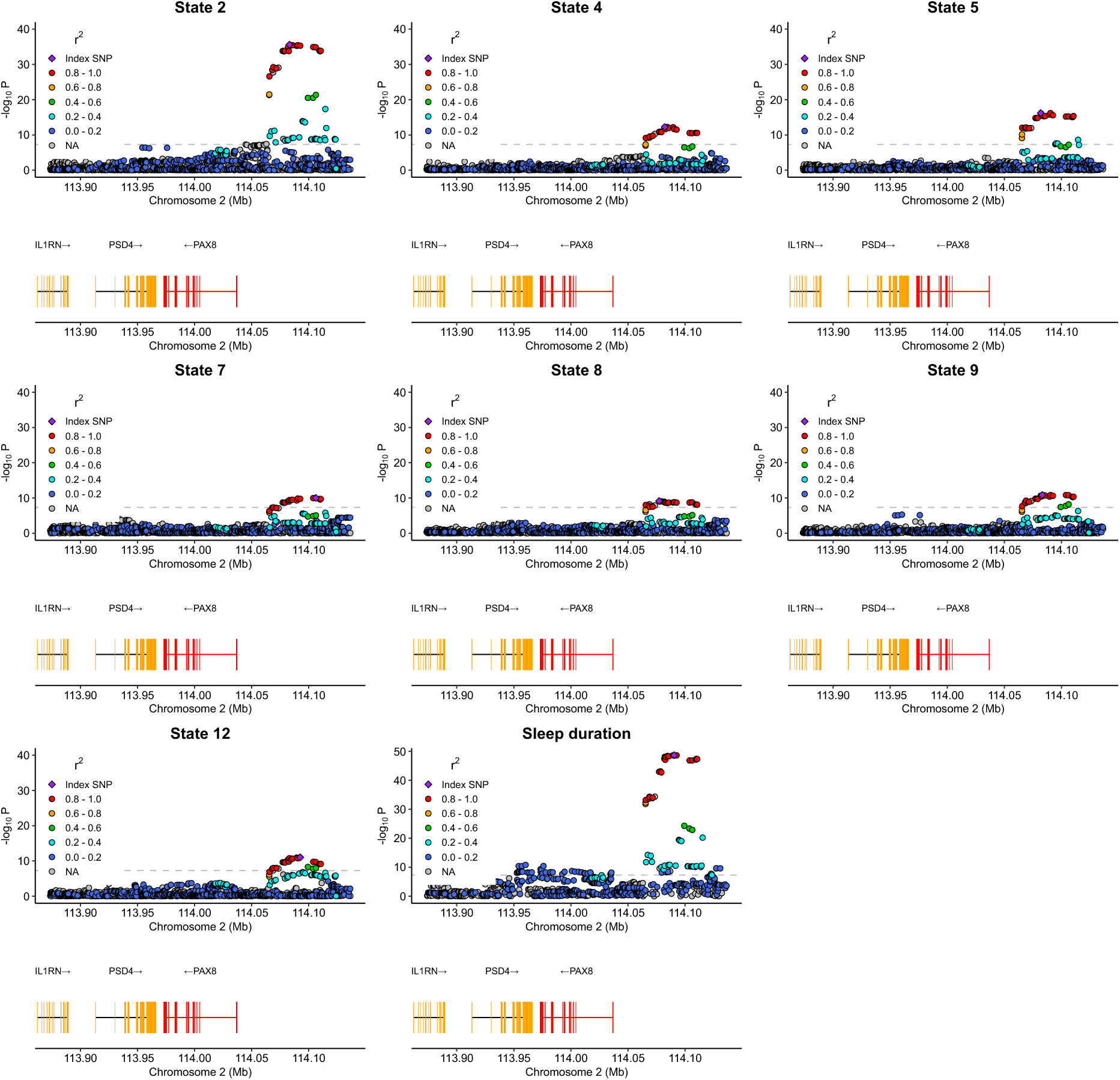
Shared genetic signals near **PAX8** on chromosome 2 between brain states (2, 4, 5, 7, 8, 9 and 12) and sleep duration (PP.H4.abf > 0.95).

**Extended Data Fig. 7.**
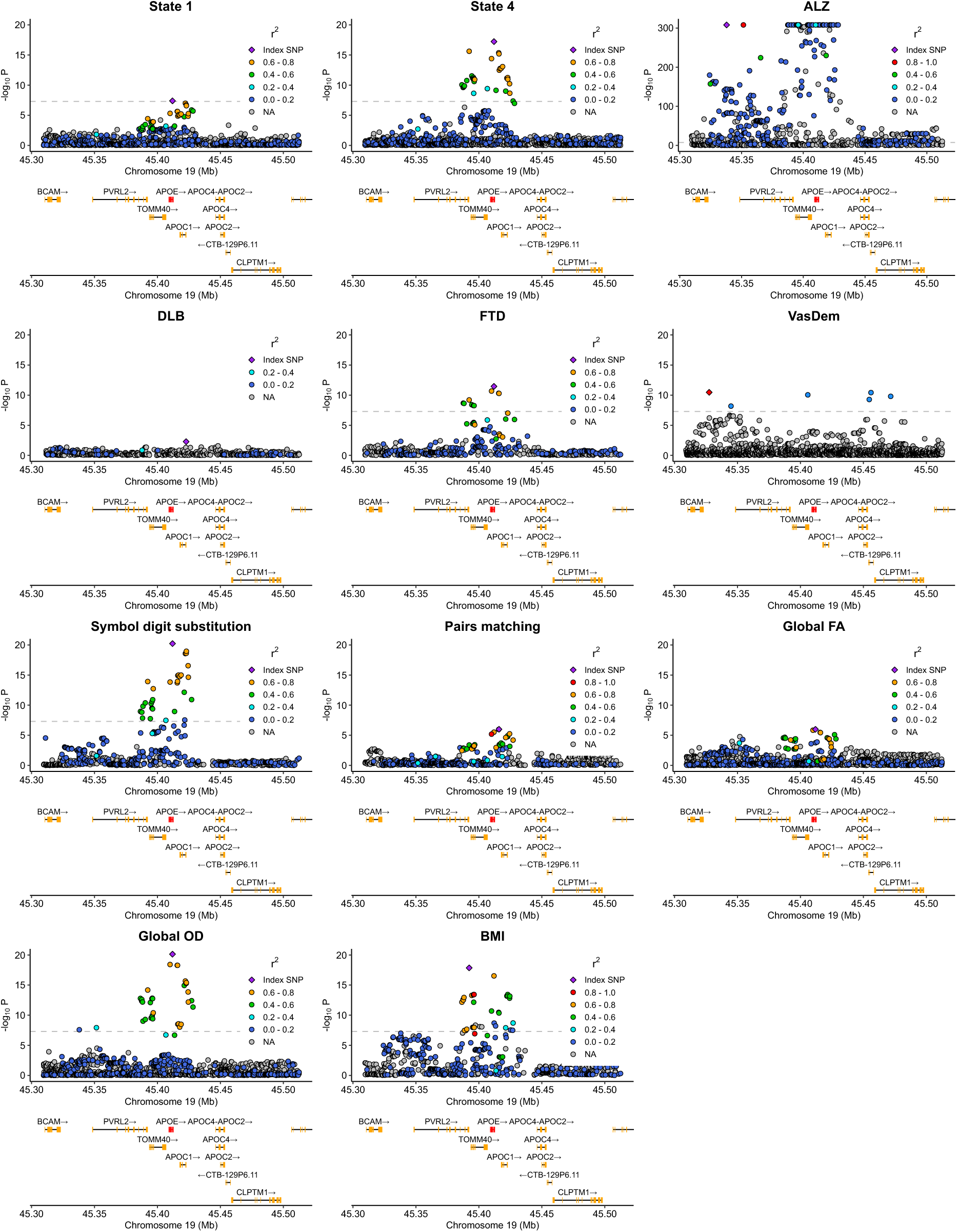
Shared genetic signals within **APOE** on chromosome 19 between brain states (1 and 4) and relevant phenotypes (PP.H4.abf > 0.95).

**Extended Data Fig. 8.**
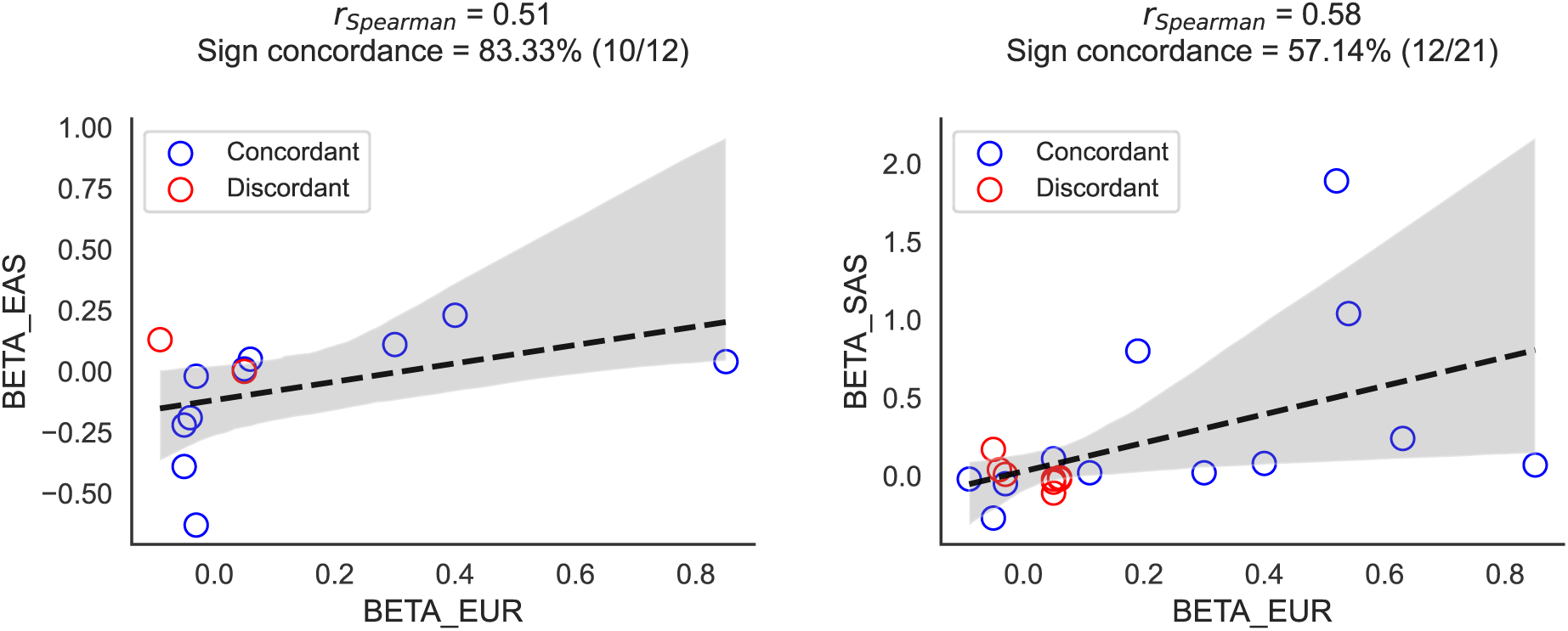
Cross-ancestry replication of lead SNPs identified in the GWAS of fractional occupancy of brain states in the UK Biobank. Shown are the sign concordance and correlation of beta estimates across European (EUR), East Asian (EAS), and South Asian (SAS) samples.

